# Longitudinal health survey of women from Venezuela in Colombia (ELSA-VENCOL): first report

**DOI:** 10.1101/2022.08.26.22279267

**Authors:** Jorge Acosta-Reyes, Laura Juliana Bonilla-Tinoco, Melissa Aguirre, Maylen Rojas-Botero, Luis Ángel Anillo, David Alejandro Rodríguez, Lida Yoana Cifuentes, Iván Jiménez, Luisa Fernanda León, Ietza Bojorquez-Chapela, Julián Alfredo Fernández-Niño

## Abstract

**Background:** Colombia is currently the world’s main recipient country for Venezuelan migrants, and women represent a high proportion of them. This article presents the first report of a cohort of Venezuelan migrant women entering Colombia through Cúcuta and its metropolitan area (the main land entry point to this country). The study aimed to describe the health status and access to healthcare services among Venezuelan migrant women in Colombia with irregular migration status, and to analyze changes in those conditions at a one-month follow-up.

**Methods:** A longitudinal cohort study of Venezuelan migrant women, 18 to 45 years, who entered Colombia with an irregular migration status, was carried out in 2021. Study participants were recruited in temporary shelters, transit points, and migrant settlements in Cúcuta and the metropolitan area. At baseline, we administered a structured questionnaire including sociodemographic characteristics, migration history, health history, access to health services, sexual and reproductive health, practice of early detection of cervical cancer and breast cancer, food insecurity, and depressive symptoms. The women were again contacted by phone one month later, between March and July 2021, and a second questionnaire was applied.

**Results:** A total of 2,298 women were included in the baseline measurement and 56.4% could be contacted again at the one-month follow-up. A significant increase was found in the percentage of women who had a health problem during the past month (from 23.1% to 31.4%; p<0.01); as well as in the share who reported moderate, severe, or extreme difficulty working or performing daily chores (from 5.5% to 11.0%; p= 0.03) and who rated their health as fair (from 13.0% to 31.2%; p<0.01). Meanwhile, the percentage of women with depressive symptoms decreased from 80.5% to 71.2% (p<0.01).

**Conclusion:** This report is a starting point for the longitudinal follow-up of the cohort, which will allow us to better understand how the health status of Venezuelan women changes during the migration flow in Colombia.

## Background

Since 2015, the social, political, and economic conditions in Venezuela have led to one of the largest international human mobility processes in the history of the region, a process that intensified in 2017 when the migration flow increased [1]. By November 2021, estimates showed roughly 6.04 million Venezuelan refugees and migrants worldwide [2]. This process, characterized as a south-south migration [3], has involved significant female participation, a phenomenon known as the feminization of the migration [4]. This contrasts with other well-known economic migration phenomena, which have historically been male-dominated.

With 1.84 million Venezuelan migrants in Colombia as of May 2022, the country is currently the world’s main recipient of Venezuelan migration [2]. In 2021, 81.3% of Venezuelan migrants in this country had an irregular migration status or were in the process of regularization, and 49.0% were women [5]. Although this is a diverse population, many of the migrants in the latest wave have been identified as having a great degree of socioeconomic vulnerability during the process of social insertion into the receiving community. Unlike the socioeconomic profile of migrants in the waves prior to 2017, this latest one has been dominated by low- and middle-income migrants with an irregular migration status who opted to migrate by land to countries near Venezuela, such as Colombia, which shares an 1,800 km border with Venezuela [1].

Among the challenges faced by migrants are the health risks and needs associated with the entire migration process, which stem from a combination of the conditions of departure, transit, and destination [6]. Since this is a mostly young population, the prevalence of chronic disease is relatively low. However, problems with access to health services can result in their becoming ill more frequently, which affects their wellbeing and at the same time can seriously impact health services. In the case of Colombia, it has been consistently reported that Venezuelan migrants are predominantly affected by skin, respiratory, gastrointestinal, immunopreventable, vector-borne, and infectious diseases (e.g., HIV/AIDS, tuberculosis, and sexually transmitted diseases), as well as by mental health disorders, gender-based violence, and health problems and needs related to pregnancy, childbirth, and the puerperium [7–10].

In addition, many migrants have encountered barriers to effective access to health services when they need them, even those that they should be able to access according to current regulations. This is due to having little knowledge about their rights and to barriers in the access routes to health services. While Colombia’s health sector has had a plan for migration [11] since 2018, most migrants with irregular immigration status can only access health services in emergencies or for public health interventions (such as vaccination). One exception is prenatal care and childbirth for pregnant women, which in theory is guaranteed regardless of migration status [11,12].

Meanwhile, migrants with a regular status can enroll in the general social security health system and access health services under the same conditions as Colombians in the country. Thus, regularization processes have helped to increase access to health insurance, which has become even more available through the recently adopted Temporary Protection Permit (PPT in Spanish), a measure that enables the regularization of all migrants who entered Colombia before January 2021. However, while regularization is a necessary condition for health insurance, it is not sufficient, and does not include other groups of migrants who do not intend to remain in the country. It also does not prevent migrants from encountering other barriers to access.

Regarding the Venezuelan migratory process in Colombia, it has been reported that migrant women use outpatient, emergency, and hospitalization services more than men, and represented nearly 70% of migrants who were treated during the period 2017-2019 [8]. Migrants 20 to 29 years of age were the group that used health services the most. In terms of the causes for seeking health services, pregnancy, childbirth, and puerperium were always among the three main reasons [8]. On the other hand, a study conducted in 2020 found that only 62.1% of pregnant Venezuelan migrants received prenatal checkups (PNC), and an analysis by migration status found that only 50.6% of migrants with irregular migration status received PNC compared to 79.5% for migrants with regular status [13].

The Colombian government has made considerable efforts to expand the supply and coverage of health services to facilitate access to health services for Venezuelan migrants, as well as to keep records of the medical care that they receive [12]. Consequently, some of the existing information systems have been modified and new ones have been created for recording the medical care received by this population, as well as for vital statistics and epidemiological surveillance.

While these information systems have been important for monitoring the health needs of Venezuelan migrants and their use of health services, as well as for guiding public health and public policy decisions, they have some limitations. There is the fact that the health information that is obtained largely depends on access to health services, which is low among migrants. The coverage is also limited and there are lags and problems with the quality of the health information on migrants [14]. In addition, the scope of these data is limited, as they do not enable the use of a multidimensional approach to migrant health, and they are insufficient for evaluating and monitoring migrant health at the population level. Moreover, a system is needed that enables identifying the health situation and needs, as well as access to health services, especially for Venezuelan migrant women since they not only represent roughly half of the migrant population but are of reproductive age and have historically been the group that has used certain health services the most. Although the Department of National Statistics (DANE in Spanish) and several organizations have gone to great lengths to conduct population-based surveys, these tend to be limited to only a few dimensions. They are also cross-sectional and have mainly focused on migrants who have already settled in the country and intend to stay.

The generation of primary source information that includes individuals regardless of their having contact with health services can provide a better understanding of the health situation of Venezuelan migrant women. Thus, the objective of this research was to describe the health status and access to health services among Venezuelan migrant women in Colombia and the changes observed at a one-month follow-up. This work presents the first report of a cohort of Venezuelan migrant women with an irregular migration status who entered Colombia through Cúcuta and its metropolitan area.

## Methods

Requests for access to data could be sent to the Migration and Health Program of the International Organization for Migration in Colombia (David Rodríguez).

### Study design and population

ELSA-VENCOL (in Spanish: *Estudio longitudinal de Salud de las mujeres Venezolanas en Colombia*) is one of the first longitudinal study among Venezuelan migrants carried out in Colombia. This was a longitudinal cohort study of Venezuelan migrant women with an irregular migration status who entered Colombia through Cúcuta and its metropolitan area, with a cross-sectional baseline measurement and a follow-up one month after. New follow-ups are expected to be done in the future.

#### Selection criteria, recruitment, and follow-up

Cúcuta and its metropolitan area is the main land entry point to Colombia. This is a region of high pendular flow (more than 35,000 people crossed per day before the pandemic), the trans-border movement of people to obtain goods and services in one side of the border, coming back to their country of residence after just a few hours or days. It is also the point where “walkers” (*caminantes*, a term used for Venezuelan migrants moving on foot) enter the country before moving on to the larger cities or continuing to the south of the continent. The flow in this region is thus mixed, including various types of migration. Although the flow decreased during the pandemic, since there was an administrative closure until the end of 2021, the crossing of people continued, mainly irregularly through informal crossings (called “*trochas*” in Spanish).

This survey included women who: 1) were between 18 and 45 years of age; 2) had entered Colombia through Cúcuta (border city with Venezuela) or its metropolitan area (Los Patios, Puerto Santander or Villa del Rosario) at any time during the baseline measurement period; 3) did not have Colombian nationality; 4) expressed the intention to remain in Colombia for at least one year; 5) were not “circular” migrants (see definition below); and 6) did not have their passport stamped when entering Colombia and did not have a valid Special Permit to Stay (PEP in Spanish), which meant that their migration status was irregular.

For this study, “circular” was defined as having entered Colombia two or more times during the month prior to the first interview for reasons other than tourism, vacations, family visits or business trips. That is, the study excluded persons who in the previous month had entered Colombia frequently for reasons of work, study or in search of services, among other purposes, since this population is considered to be part of the cross-border living dynamics rather than a migrant population.

The baseline interview was conducted face-to-face between February 15 and May 25, 2021, at various locations in the cities of Cúcuta, Los Patios and Villa del Rosario, and lasted an average of 30 minutes. In this interview, the telephone numbers of all participants were obtained, and they were informed that they would be contacted again one month later. For the follow-up interview, the women were contacted by telephone between March 16 and July 6, 2021. The second contact was attempted beginning one month after the initial interview, at different times over a period of two weeks and on different days of the week. When there was no response after this period the participant was considered to be lost to follow-up. The average telephone call lasted 25 minutes.

#### Sampling

Given the lack of a clearly defined sampling frame, a non-probability sampling method with snowball expansion was used. Interviewers went to temporary shelters and settlements where Venezuelan migrants were living and invited all migrants who were there during the fieldwork period to participate. Participants were also recruited on a road with a high flow of migrants, and all Venezuelan women walking past that point were invited to participate.

### Data collection instruments and study variables

A standardized questionnaire with 161 questions was administered to assess the following dimensions of health status and access to health services: health history and perceived morbidity, effective access to health services, sexual and reproductive health, early detection of cervical and breast cancer, food insecurity and depressive symptoms.

The participants were asked about self-perceived health, health problems or conditions over the past month, self-reported difficulty performing daily chores or work activities over the last six months (with a 6-item Likert scale ranging from none to extreme), medical diagnoses of diseases (presence or absence) and medication use (yes/no). These questions were extracted and adapted from the 2015 Colombia National Demographics and Health Survey (ENDS in Spanish). The presence of significant depressive symptoms was evaluated using the seven-item version of the Center for Epidemiological Studies depression scale (CES-D), with a cut-off point that was validated for the Colombian population [score greater than or equal to 8] [15,16].

Sociodemographic information was also collected (age, marital status, whether they were accompanied by others, number of people accompanying them, self-perceived race, educational level, sleeping place, number of people in the household, head of household, income, subsidies received) and on the migration process (country of residence, reason for migration, reason for entering Colombia through Cucuta, means of transportation, living in Colombia previously). The questions on sociodemographic conditions were adapted from the ENDS, and other questions were included that were adapted from the 2018 National Population and Housing Census of Colombia and the 2007 Study on Global AGEing and Adult Health (SAGE) questionnaire.

All surveys were reviewed by experts on the topics and evaluated with a pilot test consisting of 30 interviews with 30 migrants during the first two days of fieldwork (February 15 and 16, 2021), which allowed the questions to be adapted and modified. This pilot test demonstrated that the instruments and the items included performed well.

Similarly, a standardized questionnaire with 154 questions was used for the follow-up, which evaluated the same topics as those in the initial questionnaire, with the addition of questions on work history in Colombia. The follow-up questionnaire had fewer questions since those that were no longer relevant because it was not the first contact with the interviewers were eliminated. In addition, emphasis was placed on changes in conditions over the past month. Both interviews were conducted by previously trained interviewers.

### Statistical analysis

A descriptive analysis of the baseline data was performed using measures of central tendency and dispersion for quantitative variables and percentages for categorical variables. To assess changes between baseline and follow-up, bivariate analyses were performed with the McNemar test for dichotomous categorical variables and the marginal homogeneity test (Stuart-Maxwell) for nominal-polytomous variables. Ordinal variables were treated as polytomous categorical variables since we did not want to evaluate the correlation between the first and second measurements but rather the percentage change between the two measurements. For this reason, they were also analyzed using the marginal homogeneity test. In the case of quantitative variables, they were first evaluated using graphical and numerical methods (Shapiro-Wilk test) to identify whether they followed a normal distribution, and if they did not then the Wilcoxon test was used to compare the data from the two measurements.

To assess potential bias from loss to follow-up, the baseline characteristics of the participants with and without follow-up were compared using the chi-square test for categorical variables (and Fisher’s exact test in cases where the sample was small) and the Mann-Whitney U test for quantitative variables.

All analyses were performed using Stata® v14 with a significance level of 0.05 indicating statistically significant changes.

### Ethics statement

This study was reviewed and approved by the Ethics Committee of the Universidad del Norte. All participants gave informed consent to be included and their data were kept strictly confidential. Telephone numbers were provided by the women after the study was explained and informed consent was obtained. Informed consent was again obtained at the time of follow-up. During both interviews it was made clear to the women that their participation was free and voluntary, that it would not affect any social benefits that they may receive and that sharing the information would not pose any risk. Personal information that could lead to individual identification of the participants was only known and handled by the research team and was omitted from the final databases.

## Results

A total of 2,298 Venezuelan migrant women were surveyed at baseline and 1,297 were surveyed at one-month follow-up (56.4% response rate).

Statistically significant differences were found in the baseline measurements between women with and without follow-up, specifically, those with follow-up had a higher educational level, had been in Colombia longer, and had more contact with Colombian health services. They also had a slightly smaller percentage of significant depressive symptoms (Table S1).

### Socioeconomic and migration flow conditions

In the baseline survey, the median age was 28 years (IQR: 23-35), 57.5% of the participants were married or in a free union, 59.4% were the head of household, 97.5% did not identify as belonging to any ethnic group and 50.4% had attained only secondary school education. In addition, 34.8% indicated that they would sleep on the street that night. The median number of household members in Colombia was 4 (IQR: 3-5), 72.4% of the households had only one person earning an income, 90.9% had a household with a monthly income of less than one minimum wage ($1,000,000 Colombian pesos per month, which is equivalent to approximately $259 USD) and only 0.9% received economic assistance or subsidies (Table 1).

**Table 1.**
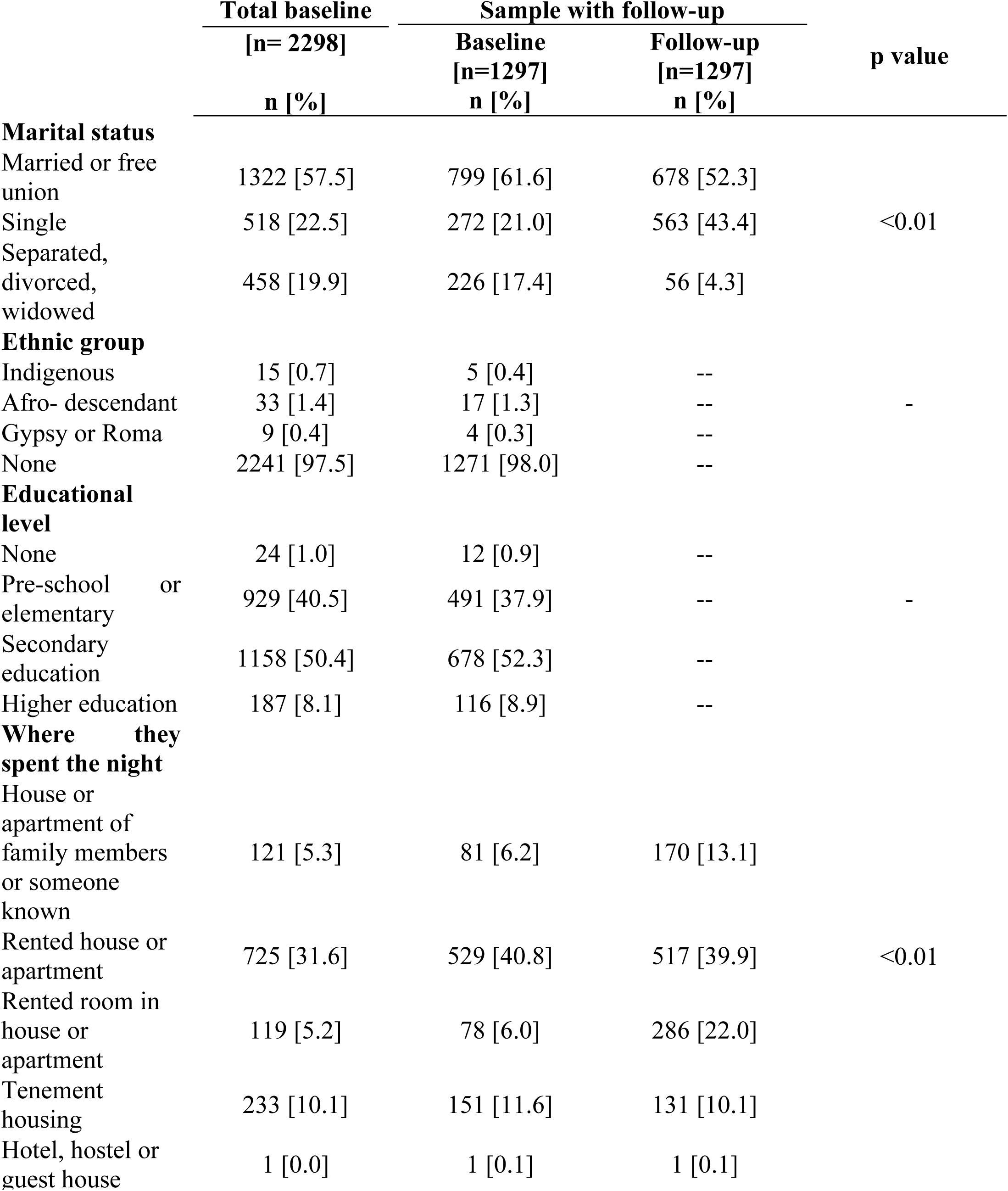
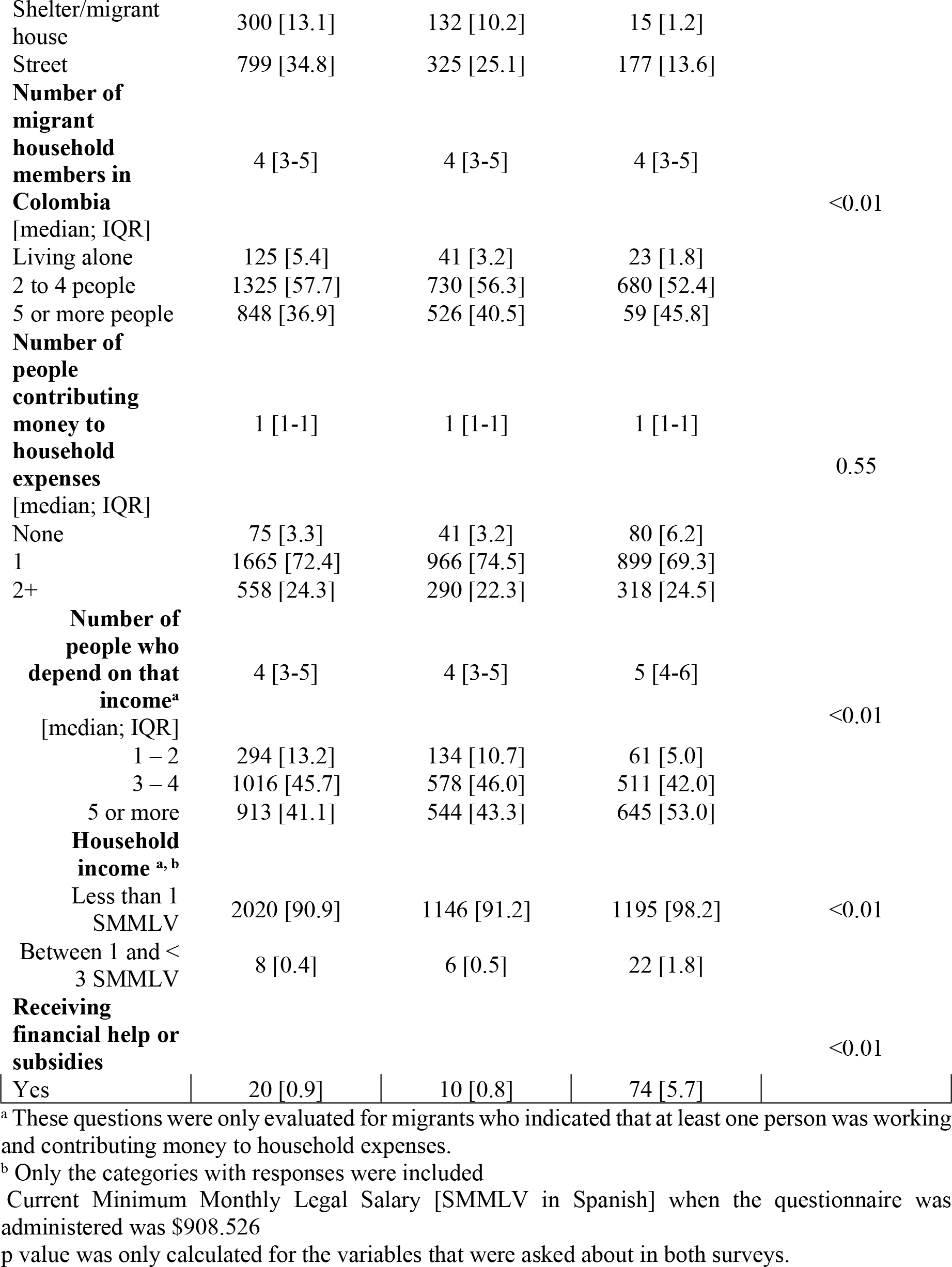
Sociodemographic data of the migrant women with irregular migration status participating in ELSA-VENCOL, 2021.

A decrease between baseline and follow-up was found in the percentage of migrants sleeping on the street, while an increase was found in the percentage of those who contributed money to the household and in participants whose households received economic assistance (Table 1). At follow-up, only 16.3% (n= 212) of the participants had initiated the process to regularize their migration status. The documents that were most frequently obtained were the PPT (n= 143; 67.5%) the PEP (34; 16.0%) and the laissez-passer (n= 35; 16.5%).

### Health status and access to health services

Regarding health conditions, at baseline 23.1% of the participants had presented a health problem or condition in the past month and 29.5% in the past 6 months, and 14.4% evaluated their health as fair or poor. The most frequent medical diagnosis was vector-borne disease (18.7%) and 80.5% of the migrants had clinically significant depressive symptoms at the time of the interview. Of the participants who needed medications, 36.4% did not take them at the time of the interview, primarily because of their cost.

At follow-up, considerably more women reported having had a health problem in the past month, with the figure rising from 23.1% to 31.4%. This increase was more notable among those who were interviewed one month or less after arriving in Colombia, 28% of whom reported a health problem in the past month, compared to those who were interviewed after being in the country for over one year, 24% of whom reported having a health problem (Table 4). Similarly, there was a marked increase in the percentage of participants who reported moderate, severe, or extreme difficulty working or performing daily chores (from 5. 5% to 11.0%) and who rated their health as fair (from 13.0% to 31.2%). Conversely, the percentage of women with depressive symptoms decreased from 80.5% to 71.2%. Table 2 shows all the results related to the health status and access to services of the participants.

**Table 2.**
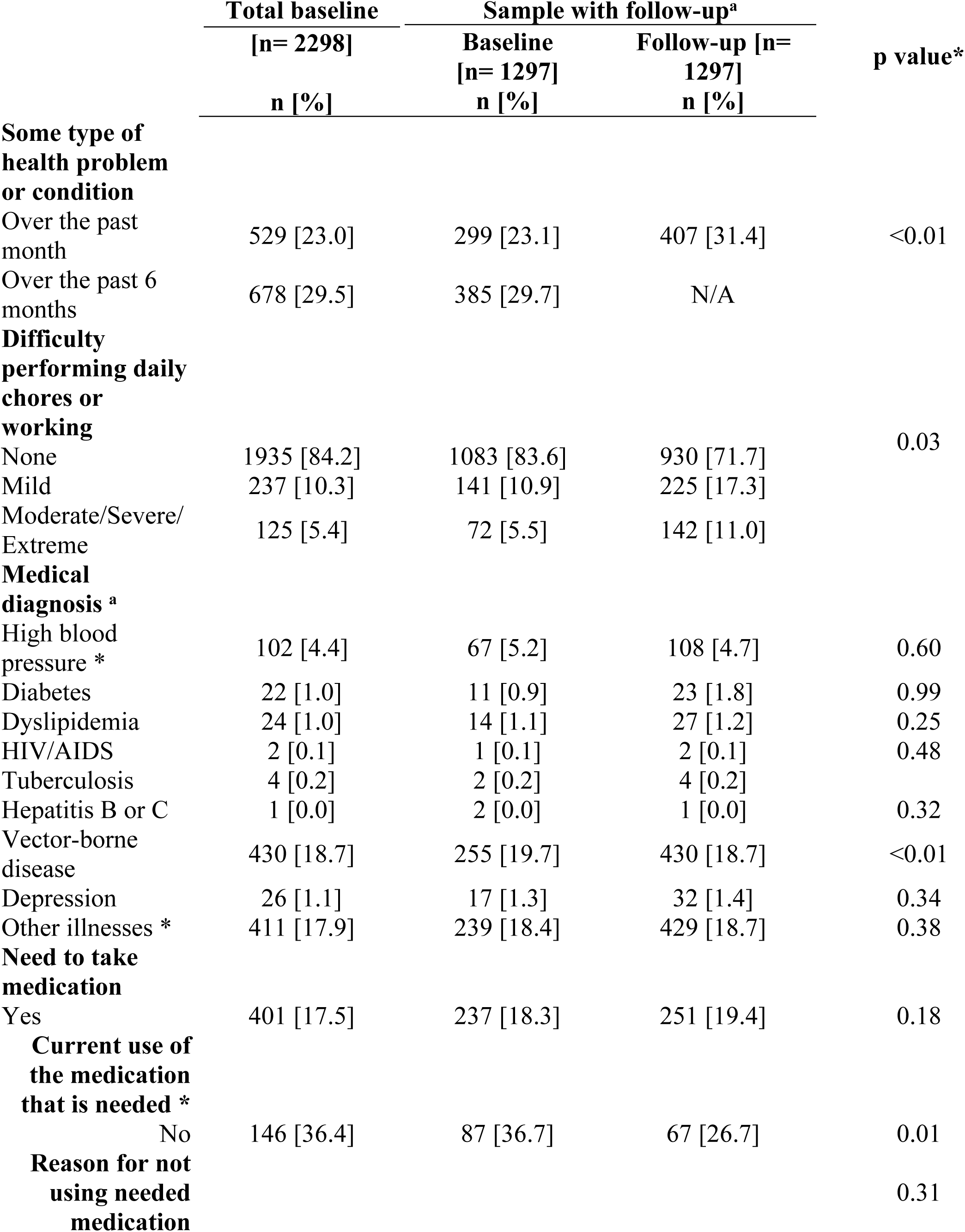
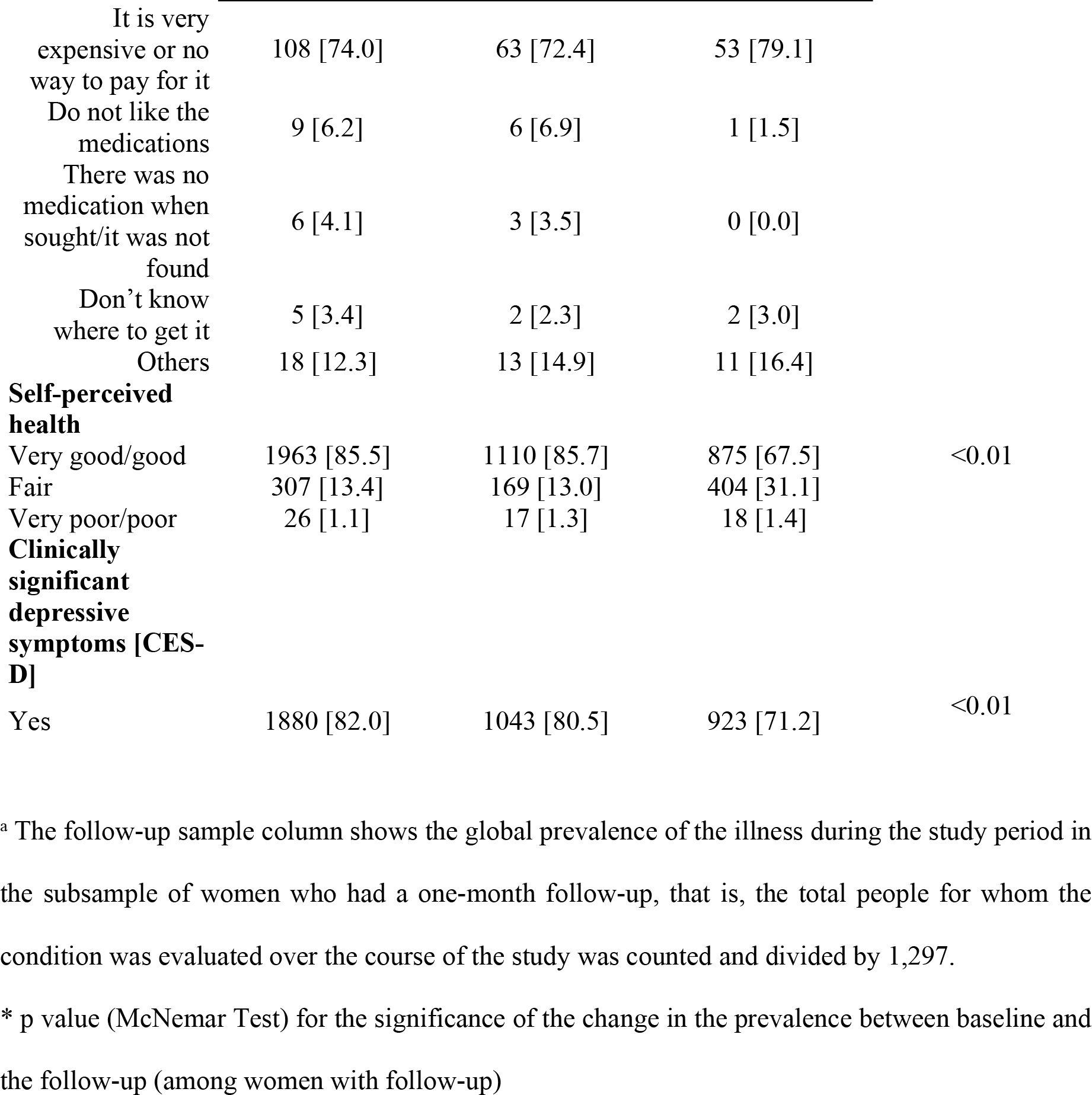
Health status, access to healthcare services and self-perceived health among the migrant women with an irregular migration status participating in ELSA-VENCOL, 2021

The differences found between baseline and one-month follow-up remained the same when categorized by length of time in Colombia: <1 month, 1 month to 1 year and >1 year (Table S2).

## Discussion

The present investigation sought to analyze the health status and access to health services of Venezuelan migrant women with irregular migration status at baseline and one-month follow-up. ELSA-VENCOL) is one of the first longitudinal study among Venezuelan migrants carried out in Colombia.

The baseline information showed that roughly one-fifth of the participants had had a health problem during the previous month. However, the prevalence of a prior diagnosis of chronic diseases was low, indicating that this was a relatively healthy population. At the same time, the prevalence of significant depressive symptoms was extremely high [82% at baseline], which likely reflects the psychosocial impact of the conditions associated with migration [17,18]. Research in a similar population (in-transit migrants in Mexico) has shown that women are more likely than men to experience common mental disorders in this type of context [19].

In this regard, it is interesting that over the follow-up period the percentage of women who reported having had some health problem or condition in the past month increased while the prevalence of significant depressive symptoms decreased. When also considering the improvement in socioeconomic factors (a higher percentage who had stable living conditions such as rented houses or rooms, received subsidies or had contact with health services), these results seem to indicate that more time in Colombia is associated with a more stable situation, which in turn is accompanied by an improved emotional state. However, since at least one study has shown that the effect of socioeconomic stressors on mental health during the resettlement process of displaced populations differ between men and women, further research to explore this hypothesis is required [20].

The increase in the prevalence of reported health problems is an interesting finding. There are very few longitudinal studies of similar populations (i.e., south-south mixed migrant flows with a combination of in-transit and resettling persons), but at least one cohort study of Bosnian refugees in Croatia found that, at a three-year follow-up, their health status had improved. Health improvement would be expected since migrants tend to experience less socioeconomic risk factors for health problems (e.g. poverty, insecure housing) over time. However, it is possible that the time frame of our study did not capture this [21]. Instead, having had more access to services at follow up could make women more likely to perceive when they had a health problem, rather than dismiss it out of hand when unable to access treatment. In addition, the slight increase in the prevalence of some of the diseases evaluated could correspond to better detection of the disease in the study population, that is, to an increase in the detection of the pathology [possibly due to greater access to health services] rather than to a real increase in the disease. While given the data in this study, these interpretations are speculative, nevertheless, it would be interesting to explore them further in the future. In addition to health-related factors, it is striking that only 16.3% of the participants who were followed had sought to regularize their migration status. Given this insufficient level of regularization, it would be worthwhile to disseminate more information and strengthen education about the process of regularizing their status in Colombia, especially in the framework of the PPT. This would dispel the myths about this migration and clarify the benefits obtained as well as those that are not lost. Regularization is also important because it directly impacts the health conditions of migrants and the health services that are available to them.

The limitations of this study include having obtained more follow-up with the subsample of participants who were already settled and had been living in Colombia for a longer period when joining the study (baseline survey). Follow-up information was obtained from only 28% of those who had been in the country for one month or less at the time of the baseline survey, compared to 55% of those who had been in Colombia for more than one month and up to one year and 74% of those who had been in Colombia for more than one year. However, the differences were not statistically significant for most of the variables. The conclusions obtained may not be fully generalizable to newly arrived migrants and those who are still in transit. In addition, due to its very nature, this survey is not exempt from the possibility of information bias. Furthermore, the response rate was 56% at follow-up. A sensitivity analysis of the population that was actually contacted one month after the initial interview indicated that the variables that had significant differences, which can guide additional analyses or follow-up studies, characterized the subsample that could be followed. This needs to be considered when making practical decisions based on the data collected. In addition, there is the possibility of information bias because it was detected that some migrants gave erroneous answers thinking that they would receive a benefit. This could not be corrected for participants without follow-up. The strengths of this study include 1) its longitudinal design, 2) the population of women given their specific health needs and that they currently represent 49% of the Venezuelan migrant population in Colombia [4], and 3) the sample size, which provided sufficient statistical power for analyzing the data. This type of analysis could help to adapt interventions so that they are more specific to the target population group.

## Conclusion

This study was able to collect a large amount of information. It is hoped that this data will be used to generate research hypotheses for subsequent studies to investigate particular relationships or associations between variables, or to provide explanations for some of the findings. It is also hoped that this study will not only help to guide the design of public policies aimed at facilitating the insertion of Venezuelan migrants into Colombian society and improving their quality of life, but that it will also serve as a first step towards beginning to close the gap in knowledge that exists with respect to multidimensional information on the health status and access to healthcare services of Venezuelan migrants in Colombia.

## Data Availability

The database of this survey is administered by the Migration and Health program of the International Organization for Migration (IOM) in Colombia. This information is reserved and cannot be made public, in accordance with existing legal frameworks, and
permission from IOM and the United States Agency for International Development (USAID) is required for its use by other researchers. Therefore, the data cannot be made publicly available. However, data access requests may be sent through David Rodriguez via email who will process them in accordance with the organization's rules and current agreement

## Acknowledgments

This publication was made possible thanks to the generous support of the people of the United States through the Community Stabilization Activities (CSA) of the U.S. Agency for International Development (USAID), the Universidad del Norte de Barranquilla and the International Organization for Migration (IOM). We thank the health authorities of Cucuta and Villa del Rosario, the local migrant organizations, and all the women who participated in this study.

